# Rapid Viral Expansion Beyond the Amazon Basin: Increased Epidemic Activity of Oropouche Virus Across the Americas

**DOI:** 10.1101/2024.08.02.24311415

**Authors:** Felipe Campos de Melo Iani, Felicidade Mota Pereira, Elaine Cristina de Oliveira, Janete Taynã Nascimento Rodrigues, Mariza Hoffmann Machado, Vagner Fonseca, Talita Emile Ribeiro Adelino, Natália Rocha Guimarães, Luiz Marcelo Ribeiro Tomé, Marcela Kelly Astete Gómez, Vanessa Brandão Nardy, Adriana Aparecida Ribeiro, Alexander Rosewell, Álvaro Gil A. Ferreira, Arabela Leal e Silva de Mello, Brenda Machado Moura Fernandes, Carlos Frederico Campelo de Albuquerque, Dejanira dos Santos Pereira, Eline Carvalho Pimentel, Fábio Guilherme Mesquita Lima, Fernanda Viana Moreira Silva, Glauco de Carvalho Pereira, Houriiyah Tegally, Júlia Deffune Profeta Cidin Almeida, Keldenn Melo Farias Moreno, Klaucia Rodrigues Vasconcelos, Leandro Cavalcante Santos, Lívia Cristina Machado Silva, Livia C. V. Frutuoso, Ludmila Oliveira Lamounier, Mariana Araújo Costa, Marília Santini de Oliveira, Marlei Pickler Dediasi dos Anjos, Massimo Ciccozzi, Maurício Teixeira Lima, Maira Alves Pereira, Marília Lima Cruz Rocha, Paulo Eduardo de Souza da Silva, Peter Rabinowitz, Priscila Souza de Almeida, Richard Lessells, Ricardo T. Gazzinelli, Rivaldo Venâncio da Cunha, Sabrina Gonçalves, Sara Cândida Ferreira dos Santos, Senele Ana de Alcântara Belettini, Silvia Helena Sousa Pietra Pedroso, Sofia Isabel Rótulo Araújo, Stephanni Figueiredo da Silva, Julio Croda, Ethel Maciel, Wes Van Voorhis, Darren P. Martin, Edward C. Holmes, Tulio de Oliveira, José Lourenço, Luiz Carlos Junior Alcantara, Marta Giovanetti

**Affiliations:** Central Public Health Laboratory of the State of Minas Gerais, Ezequiel Dias Foundation, Brazil; Central Public Health Laboratory of the State of Bahia, Brazil; Central Public Health Laboratory of the State of Mato Grosso, Brazil; Central Public Health Laboratory of the State of Acre, Brazil; Central Public Health Laboratory of the State of Santa Catarina, Brazil; Department of Exact and Earth Sciences, University of the State of Bahia, Salvador, Brazil; Centre for Epidemic Response and Innovation (CERI), School of Data Science and Computational Thinking, Stellenbosch; René Rachou Institute, Oswaldo Cruz Foundation, Belo Horizonte, Brazil University; Stellenbosch, South Africa; Organização Pan-Americana da Saúde, Organização Mundial da Saúde, Brazil; Institute of Biological Sciences, Federal University of Minas Gerais, Belo Horizonte, Brazil; Coordenadora-Geral de Vigilância de Arboviroses, Brazilian Ministry of Health, Brazil; Coordenadora-Geral de Laboratórios de Saúde Pública, Brazilian Ministry of Health, Brazil; Unit of Medical Statistics and Molecular Epidemiology, University Campus Bio-Medico of Rome, Italy; Environmental and Occupational Health Sciences, University of Washington, USA; KwaZulu-Natal Research Innovation and Sequencing Platform (KRISP), Nelson R Mandela School of Medicine, University of KwaZulu-Natal, Durban 4001, South Africa; Fundação Oswaldo Cruz - Minas, Laboratory of Immunopatology, Belo Horizonte, MG, Brazil; Fundação Oswaldo Cruz, Bio-Manguinhos, Rio de Janeiro, Rio de Janeiro, Brazil; Faculdade de Medicina, Universidade Federal de Mato Grosso do Sul, Campo Grande, MS, Brazil; Fundação Oswaldo Cruz - Mato Grosso do Sul, Campo Grande, MS, Brazil; Secretária de Vigilância em Saúde e Ambiente (SVSA - Ministério da Saúde), Brazil; Center for Emerging and Re-emerging Infectious Diseases (CERID), University of Washington; Computational Biology Division, Department of Integrative Biomedical Sciences, Institute of Infectious Disease and Molecular Medicine, Faculty of Health Sciences, University of Cape Town, Cape Town, South Africa; School of MedicalSciences, University of Sydney, Sydney, NSW, Australia; School for Data Science and Computational Thinking, Faculty of Science and Faculty of Medicine and Health Sciences, Stellenbosch University, South Africa; Universidade Católica Portuguesa, Católica Medical School, Católica Biomedical Research Centre, Portugal; Climate amplified diseases and epidemics (CLIMADE) Europe, Portugal; Department of Sciences and Technologies for Sustainable Development and One Health, Università Campus Bio-Medico di Roma, Italy; Oswaldo Cruz Institute, Oswaldo Cruz Foundation, Minas Gerais, Brazil

**Keywords:** OROV, Brazil, genomic surveillance, Amazon basin

## Abstract

**Summary:** Oropouche virus (OROV), initially detected in Trinidad and Tobago in 1955, has been historically confined to the Amazon Basin. However, since late 2022, OROV has been reported in northern Brazil as well as urban centers in Bolivia, Colombia, Cuba, and Peru. Herein, we describe the generation of 133 new publicly available full genomes. We show how the virus evolved via genome component reassortment and how it rapidly spread across multiple states in Brazil, causing the largest outbreak ever recorded outside the Amazon basin, including the first detected deaths. This work highlights the need for heightened epidemiological and genomic surveillance and the implementation of adequate measures in order to mitigate transmission and the impacts on the population.

**Background:** Oropouche virus was first identified in 1955 in Trinidad and Tobago and later found in Brazil in 1960. Historically, it has been reported to have caused around 30 outbreaks, mostly within the Amazon Basin, where it circulates among forest animals, but also in urban areas where it is known to be transmitted by the midge *Culicoides paraensis*. Recently, Brazil has seen a surge in cases, with more than 7000 reported by mid-2024 alone.

**Methods:** In a collaboration with Central Public Health Laboratories across Brazilian regions, we integrated epidemiological metadata with genomic analyses of recently sampled cases. This initiative resulted in the generation of 133 whole genome sequences from the three genomic segments (L, M, and S) of the virus, including the first genomes obtained from regions outside the Amazon and from the first ever recorded fatal cases.

**Findings:** All of the 2024 genomes form a monophyletic group in the phylogenetic tree with sequences from the Amazon Basin sampled since 2022. Our analyses revealed a rapid north-to-south viral movement from the Amazon Basin into historically non-endemic regions. We identified 21 reassortment events, although it remains unclear if genomic evolution of the virus enabled the virus to adapt to local ecological conditions and evolve new phenotypes of public health importance.

**Interpretation:** Both the recent rapid spatial expansion and the first reported fatalities associated with Oropouche (and other outcomes under investigation) underscore the importance of enhancing surveillance for this evolving pathogen across the Region. Without any obvious changes in the human population over the past 2 years, it is possible that viral adaptation, deforestation and recent climate change, either alone or in combination, have propelled Oropouche virus beyond the Amazon Basin.

**Research in context:** *Evidence before this study:* Before this study, Oropouche virus (OROV) was known to cause periodic outbreaks primarily within the Amazon Basin. Initially identified in Trinidad and Tobago in 1955, the virus had been responsible for approximately 30 outbreaks in Latin America, mostly confined to the Amazon region. The virus typically circulates among forest animals and is transmitted to humans by the bite of the midge *Culicoides paraensis*. There has been an historical dearth of available genomic data, and so far the spread beyond the Amazon Basin has not been well-documented.

*Added value of this study:* This study provides a timely and comprehensive analysis of epidemiological and genomic data of Oropouche virus from regions outside the Amazon Basin. By generating 133 whole genome sequences from various regions across Brazil, this study reveals the movement of OROV over the past few years across Brazil, with a north-south spatial movement from regions of historic endemicity to regions with clear epidemic potential. We identify 21 reassortment events, with the possible occurrence of virus adaptation to new environments. We also report the first fatal cases of Oropouche virus infection in patients without underlying relevant comorbidities, underscoring the public health risk of future outbreaks and the need for increased awareness and surveillance.

*Implications of all the available evidence:* The rapid spread of Oropouche virus beyond the Amazon Basin into regions of Brazil more than 3500 Km distant, coupled with the identification of genome reassortment events, raises the possibility that the virus is adapting to the new environments of its increasing spatial landscape. This evolution could lead to the emergence of new viral phenotypes, with potential changes at various levels, from vector efficiency to disease outcome, raising the challenge of managing future outbreaks. This underscores the critical need for enhanced surveillance systems at national and continental levels, particularly in urban centers that appear to have been hit hard during the spatial expansion, to detect and respond to Oropouche virus outbreaks promptly.

## Introduction

Oropouche virus (OROV; *Orthobunyavirus oropoucheense*) is an arthropod-borne virus classified within the order *Bunyavirales*, family *Peribunyaviridae* and genus *Orthobunyavirus* (1). It is present as a negative-sense, single-stranded RNA genome that is divided into three segments based on size: S (small), M (medium) and L (large). These segments encode four structural proteins: the nucleocapsid, two external glycoproteins, and the RNA polymerase (2). OROV was first identified in 1955 in Oropouche, a village in Trinidad and Tobago (3). Since then, the virus has been responsible for numerous outbreaks, mostly within the Amazon Basin, where it is found among forest animals such as non-human primates, sloths, and birds (4). The midge *Culicoides paraensis* is the primary vector for human transmission. Infection with OROV causes Oropouche fever, which typically presents as fever, headache, muscle and joint pain (5). The majority of human infections present as mild to moderate disease and resolve within a week, although rare cases can lead to complications like aseptic meningoencephalitis (6).

Historically, OROV outbreaks were largely restricted to the Amazon region, with about 30 outbreaks reported in Latin America until recent years (7). However, new epidemiological data show a marked increase in Oropouche fever cases in Brazil, Cuba (N=74), Bolivia (N=356), Colombia (N=74), and Peru (N=290), with travel-related cases reported in Italy, Spain, and Germany, all originating from Cuba, highlighting that it’s becoming recognized outside the region (8, 9). Notably, Brazil alone reported a total number of more than 7,000 cases this year (up to August 2024) compared to 836 in 2023, indicating a significant rise in transmission (10-12). In 2024, Brazil also reported the first three fatal cases associated with OROV infection, raising concerns among health authorities (13). OROV has also recently appeared in historically non-endemic Brazilian regions outside the Amazon, including in the far south and, importantly, in some urban centers on the east coast (13). As is the case for other arboviruses (14), recent changes in disease ecology, such as deforestation, urbanization, human mobility and climate change, are some of the possible drivers of its recent emergence in the Amazon region (15). Local reservoir habitats belonging to non-human mammals and vectors can be disrupted, pushing them into closer contact with each other and with urban and peri-urban areas where humans can be infected. In addition, human mobility favors long distance viral movement. Additionally, ongoing changes in OROV genetic diversity may also result in changes in virulence and transmission potential (16).

Scientific and surveillance data for OROV are currently limited, with fewer than 110 peer-reviewed publications compared to thousands for other arboviruses such as Zika or Dengue (17). Reports of recent increases in OROV epidemic activity underscore the urgent need for more data and research. Some of the epidemic activity over the past decade has provided valuable insights into OROV epidemiological characteristics, as well as its potential for public health impact. To help fill current knowledge gaps in the face of recent epidemic activity in the south of Brazil we, in collaboration with several Central Public Health Laboratories, generated 133 viral genome sequences including all three segments (L, M, and S). Herein, we present genomic analysis that offers insights into OROV’s recent movement from northern to southern Brazil and its emergence in regions, within and outside the country, classically not associated with epidemic activity.

## Methods

### Ethics statement

This project was reviewed and approved by the Ethical Committee of the Federal University of Minas Gerais (CAAE: 32912820.6.1001.5149). The availability of the samples for research purposes during outbreaks of national concern is allowed by the terms of the 510/2016 Resolution of the National Ethical Committee for Research (CONEP - Comissão Nacional de Ética em Pesquisa, Ministério da Saúde) of the Brazilian Ministry of Health (BrMoH), that authorize, without the necessity of an informed consent, the use of clinical samples collected in the Brazilian Central Public Health Laboratories to accelerate knowledge building and contribute to surveillance and outbreak response. The samples processed in this study were obtained anonymously from material collected during routine arboviral diagnosis in Brazilian public health laboratories within the BrMoH network.

### Sample collection and molecular diagnostic screening

Clinical samples from patients with suspected OROV infection presenting with acute febrile illness were obtained for routine diagnostic purposes at local health services in five different Brazilian states (Minas Gerais, Bahia, Mato Grosso, Acre and Santa Catarina). Samples were collected between February and May 2024. Viral RNA was extracted from serum samples using an automated protocol, and samples were submitted to a multiplex arbovirus molecular screening by RT-qPCR, including the detection of OROV based on an assay by Naveca et al. (2017) (18). All of these samples yielded positive results only for OROV.

### cDNA synthesis and whole genome sequencing

Samples were selected for sequencing based on the CT value (≤36) and availability of clinical and epidemiological metadata, such as date of symptom onset, date of sample collection, sex, age, municipality of residence, and symptoms. For cDNA synthesis, the ProtoScript II First Strand cDNA Synthesis kit (NEB) was used following the manufacturer’s instructions. The cDNA generated was subjected to multiplex PCR sequencing using Q5 High Fidelity Hot-Start DNA Polymerase (NEB) and a set of specific primers designed by the Zibra Project (https://github.com/zibraproject/zika-pipeline/tree/master/schemes/OROV400/V1) for sequencing the complete genomes of OROV. Whole genome sequencing was performed using both MiSeq (Illumina) and MinION (Oxford Nanopore Technologies) instruments. In the first case, OROV library preparation was carried out using the KAPA HyperPlus kit (Roche), following the manufacturer’s instructions. The normalized library was loaded onto a 300-cycle MiSeq Reagent Micro Kit v2 and run on the MiSeq platform (Illumina). For nanopore sequencing, DNA library preparation was performed using the ligation sequencing kit LSK109 (Oxford Nanopore Technologies) and the native barcoding kit EXP-NBD196 (Oxford Nanopore Technologies). Sequencing libraries were loaded into an R9.4 flow cell (Oxford Nanopore Technologies).

### Generation of consensus sequences

Raw files were basecalled and demultiplexing was done using Guppy v.6.0 (Oxford Nanopore Technologies). Consensus sequences were generated by a hybrid approach using the Genome Detective online tool (https://www.genomedetective.com/) (19). The newly generated OROV sequences were deposited in GenBank and will be made available after acceptance.

### Phylogenetic analysis

Sequences of the 133 complete S, M, and L genomic segments of OROV generated in this study were combined with corresponding segments of all published full-length OROV sequences available in NCBI up to July 2024 (S=376 sequences, M=231 sequences, and L=303 sequences). The sequences from two fatal cases from the state of Bahia (Brazil) collected in March and May 2024, along with one from Mato Grosso, were also included. Sequence alignment of each segment data set was performed using MAFFT (20) and manually curated to remove artifacts using AliView (21). The full genome dataset (with segments concatenated) was checked for potential recombination and reassortment using the program, the RDP5 (using default settings except that segments were considered as linear sequences and window sizes of 16, 50, 40 and 101 were respectively used for the RDP, Chimaera, Maxchi and Bootscan recombination/reassortment detection methods; 22). Genomic regions identified by RDP5 to have been acquired by recombination and genomic segments identified by RDP5 to have been acquired by reassortment, were stripped from the full genome datasets by replacing tracts of sequence acquired through recombination/reassortment with gap characters (“-”) in the alignment file to yield a recombination and reassortment free full genome alignment. The full genome alignment and those of the individual segements were used to infer Maximum Likelihood (ML) phylogenetic trees using IQ-TREE version 2 (23) under the HKY nucleotide substitution model, which was inferred by the ModelFinder application. Branch support was assessed using the approximate likelihood-ratio test based on bootstrap and the Shimodaira–Hasegawa-like procedure with 1,000 replicates.

Three different data subsets containing only the 2022-2024 extra-Amazon sequences of S (n=162), M (n=162), and L (n=162) segments, were used to infer spatiotemporal spread patterns from continuous spatially-explicit phylogeographic reconstructions using BEAST v1.10.4 (24). Before phylogeographic analysis, the molecular clock signal in each data subset was assessed using the root-to-tip regression method available in TempEst v1.5.3 (25) following the removal of potential outliers that may violate the molecular clock assumption. We accepted temporal structure when the correlation coefficient was >0.2. We modeled the phylogenetic diffusion and spread of OROV within Brazil by analyzing localized transmission (between Brazilian regions) using a flexible relaxed random walk diffusion model (26) that accommodates branch-specific variation in rates of dispersal, with a Cauchy distribution and a jitter window size of 0.01 (27). For each sequence, latitude and longitude coordinates of the sample were considered. MCMC analyses were set up in BEAST v1.10.4, running in duplicate for 50 million interactions and sampling every 10,000 steps in the chain. Convergence for each run was assessed in Tracer v1.7.1 (effective sample size for all relevant model parameters >200) (28). Maximum clade credibility trees for each run were summarized using TreeAnnotator after discarding the initial 10% as burn-in. Finally, we used the R package seraphim (29) to extract and map spatiotemporal information embedded in the posterior trees.

To better understand the global dissemination of a specific Brazilian sublineage from 2022-2024, we expanded the data set for each segment to include genome sequences recently isolated in Peru and Italy. We then constructed time-scaled global tree topologies and performed discrete ancestral state reconstruction (of locations) using the *mugration* package extension of TreeTime under a GTR model (30). Using a custom Python script, we tracked the number of state changes by iterating over each phylogeny from the root to the external tips. We recorded state changes whenever an internal node transitioned from one country to a different country in its child node or tip(s). The timing of these transition events was documented, providing estimates for import or export events (30).

## Results

Between late 2022 and early 2024, the Brazilian states of Acre, Amazonas, Rondônia, and Roraima, located in the western Amazon region, reported a sharp increase in the incidence of OROV human cases (11). Concurrently, there was a substantial increase in the number of real-time RT-PCR tests conducted across the country, reflecting heightened screening efforts (**Figure 1a**). In 2020, initial screening efforts focused primarily on the northern region of Brazil, with the Roraima and Pará states accounting for the majority of the 238 tests conducted (31). At the same time, a few positive cases were detected in Amapá, Amazonas, Pará, Piauí, and Rondônia, signaling the early spread of OROV. By 2021, the number of tests surged to 1,466, with significant increases in states outside the Amazon basin such as Minas Gerais and Ceará. Positive cases were reported predominantly in Amapá, Pará, and Piauí (13). The trend of increased testing continued into 2022, with 588 tests conducted, marking notable expansions in Midwestern and Northeastern Brazilian states. In 2023, the screening efforts intensified further, with 5,280 tests conducted nationwide, with large testing numbers in Bahia, Goiás, Rio de Janeiro, and Tocantins, reflecting widespread concern. Positive cases were detected in multiple states, including Acre, Maranhão, Mato Grosso, Pará, Piauí, Rio Grande do Norte, Rondônia, Roraima, and Tocantins (**Figure 1a**) (13). By early 2024, screening efforts reached an unprecedented level, with 54,428 tests conducted across numerous states (**Figure 1a**). As shown in the inset panel, the positive rate nearly doubled, increasing from 0.059 in 2020 to 0.1 in 2024. The states of Espírito Santo, Minas Gerais, Bahia, and Goiás reported the highest testing numbers outside the Amazon basin (13). Within this period, cumulative positive cases were highest within Rondônia (n=1,747), Bahia (n=837), Espírito Santo (n=416), and Roraima (n=244) (**Figure 1b**).

**Figure 1.**
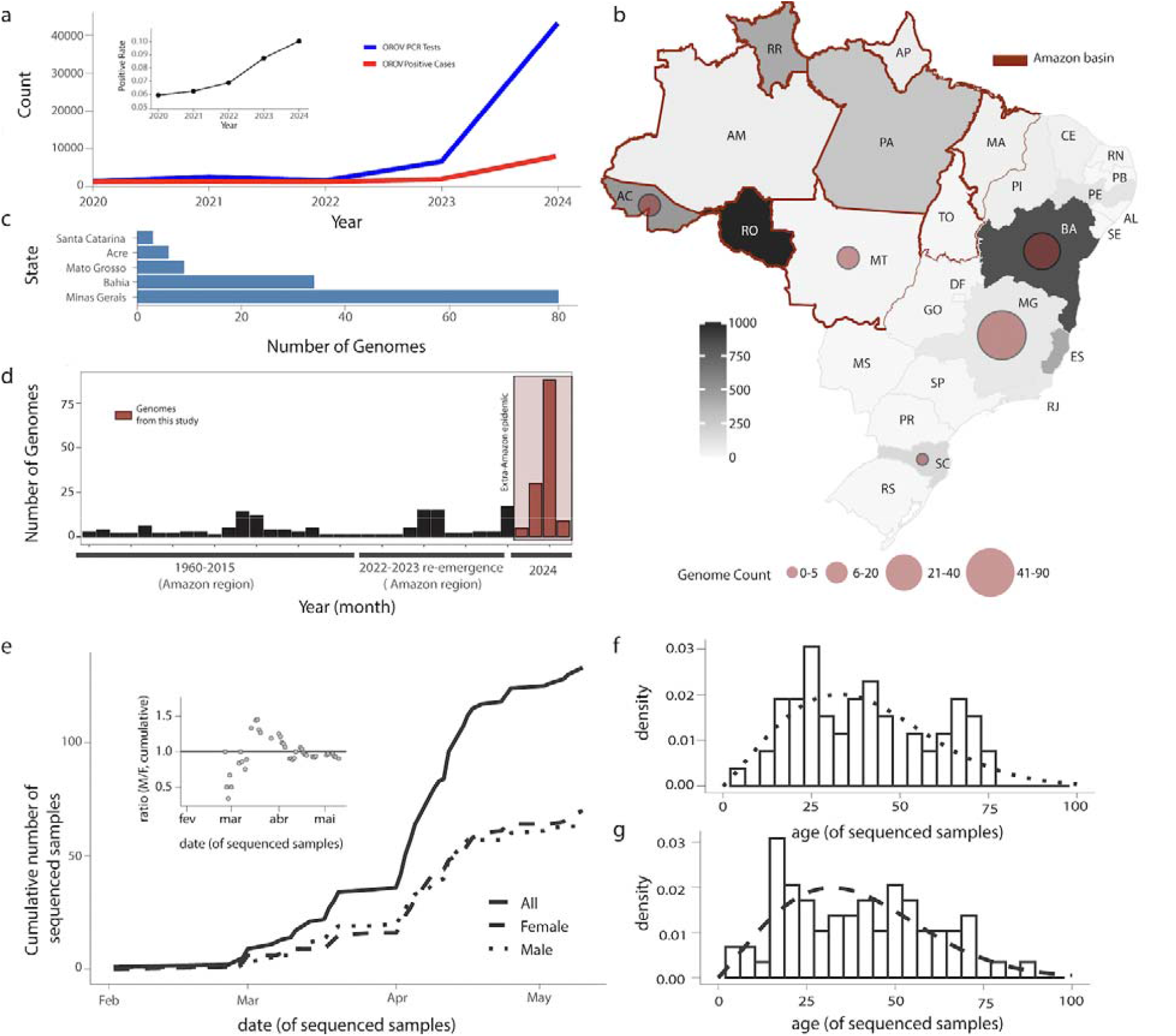
Distribution and Epidemiological Insights of OROV Clinical Cases Detected Beyond the Amazon Basin. a) Weekly notified OROV PCR tests and positive cases normalized per 100K individuals per region from 2020 to 2024. The inset panel shows the positive rate (ratio of positive cases to PCR tests) over the same period, with the y-axis scaled from 0.05 to 0.1 to highlight the variations; b) Map of Brazil showing the number of new OROV sequences by state. The color and size of the circles indicates the number of new genomes generated in this study; c) Number of new OROV genomes per state obtained in this study (Santa Catarina n=3; Acre n=6, Mato Grosso n=9; Bahia n=34; Minas Gerais n=81); d) Number of genomes generated in this study compared to the number of Brazilian OROV sequences available on the GenBank up to 31st July, 2024. Bars are months, months with 0 sampling are not shown; e) Cumulative OROV cases in absolute (full line) and by gender (male dotted, female dashed) of samples from 2024 for which gender metadata was available; the inner panel shows the gender ratio (M/F) per date; f-g) Observed (bars) and theoretical (lines) age-distributions for males (f) and females (g) of samples from 2024 for which gender metadata was available. Fitted theoretical distributions were Weibul (male: shape 2.11, scale 44.39, mean 39.15; female: shape 2.07, scale 44.22, mean 39.16).

Five Brazilian states were represented in the 133 newly generated genome sequences: Acre (N=6, northwest), Bahia (N=34, northeast), Mato Grosso (N=9, midwest), Minas Gerais (N=81, southeast) and Santa Catarina (N=3, south) (Figure 1c,a). This sampling covered a wide spatial range outside the Amazon basin (**Figure 1a**), representing the most time-intensive sampling period to date in Brazil (**Figure 1d**). The average cycle threshold (CT) for the three genes was 25, ranging from 8 to 36 (Table S1). The gender ratio associated with the genome samples was biased towards females in February-March 2024, but converged to 1 into late local autumn (**Figure 1e**). Cumulatively, 53% (n=70) were female and 47% (n=63) were male (**Table S1**), with genders showing a similar age profile (**Figure 1f-g**). Ages ranged from 1 to 89 years, with a median age of ∼39 years.

Genome sequences were obtained from all five Brazilian macro-regions (North, South, Northeast, South East and North East), revealing north-to-south viral movement across the country (**Figure 1**). The most frequent symptoms observed among the patients were fever, myalgia, and headache. These symptoms were consistently reported across multiple cases, with some patients also experiencing arthralgia. In addition to the common symptomatic presentations, our study identified three fatal cases associated with OROV infection. Notably, these fatalities occurred in young adult patients with no reported comorbidities. In all cases, the clinical course resembled severe dengue, with shock, bleeding, and extensive coagulopathy. Without the extensive lab evaluations due to the ongoing OROV outbreak in the region, these deaths would likely have been misclassified as Dengue Fever rather than OROV. The detection of these fatal cases suggests that OROV may have a broader clinical impact than previously understood, warranting further investigation into the factors contributing to severe disease outcomes. Point mutations were identified in all three fatal cases. In the M segment, amino acid changes included I to V at position 13, M to I at position 642, A to T at position 752, and R to K at position 1342. In the L segment, mutations were found at amino acid positions 857 (T to A), 1634 (K to E), and 2206 (N to D). Further in-depth analyses are required to determine whether these mutations play any role in the pathogenesis and severity of OROV infections. There was seemingly no evidence that symptoms varied with age or gender (**Figure S1**).

The sequencing procedures yielded an average coverage of 97.7% for segment S, 98.5% for segment M, and 98.32% for segment L (**Table S1**). Using a Generalized Additive Model similar to the one used for other arboviruses (30), we generated response curves for resulting sequence coverage dependent on sample CT. As expected, both observed (**Table S1**) and estimated sequence coverage were negatively correlated with CT value (**Figure 2a**). Segment M overperformed and segment S underperformed in sequence coverage independently of CT value (albeit only marginally). Segment M was the least sensitive to CT value, showing consistent sequence coverage across sample CT. The states of Bahia and Minas Gerais (∼26 and ∼61% of samples, respectively) represented extremes of model output, with consistently lower sequence coverage for Bahia and higher coverage for Minas Gerais, independently of sample CT and across all genome segments (**Figure S2**). Prolonged intervals between symptom onset and sample collection were associated with increased CT values (**Figure 2b**), suggesting a decline in viral RNA quantity and consequently sample quality over time, underscoring the critical importance of minimizing the time between symptom onset and sample collection to ensure the acquisition of high-quality sequencing data. Additionally, considering the segmented nature of the OROV genome, we investigated the potential presence of reassortment and recombination among the three different genome segments. We identified 21 reassortment events: 17 separating the S and M segments, 7 separating the S and L segments, and 11 separating the M and L segments.

**Figure 2.**
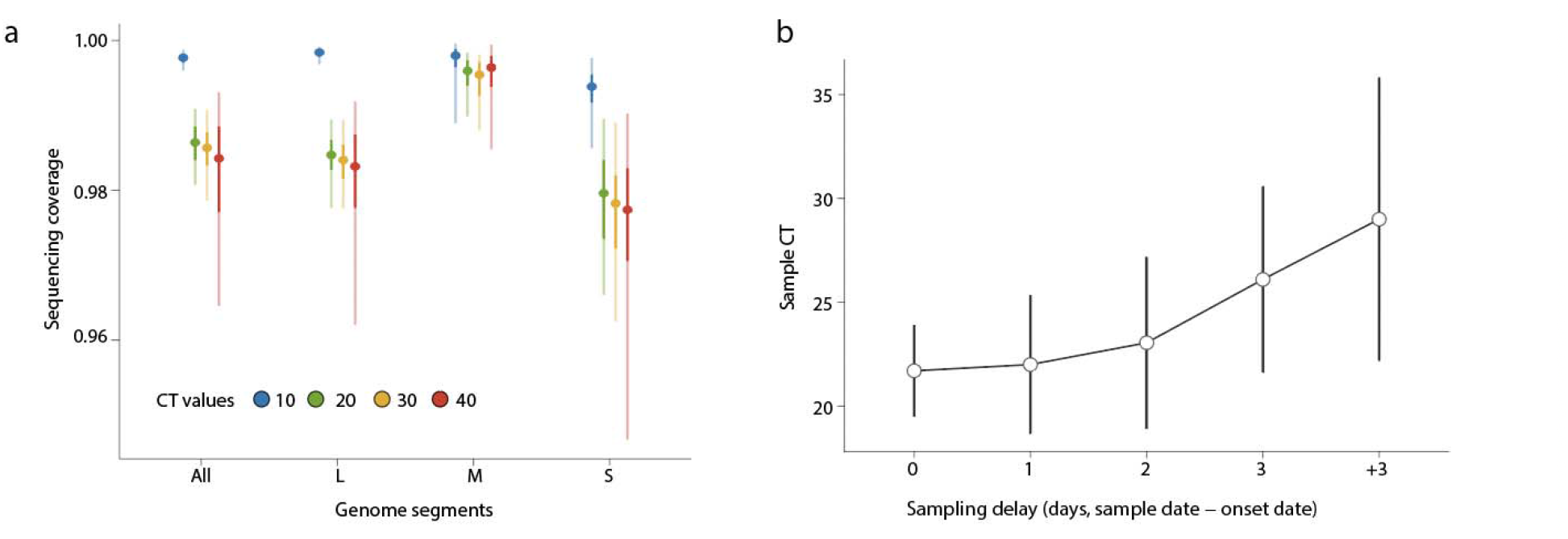
Sequencing Coverage and Sample CT Value Analysis: Model Estimates. a) Summary of estimated sequence coverage dependent on sample CT using a Generalized Additive Model. Shown are the mean (points) and 95% CI (ranges) for model estimates given selected values of CT (10, 20, 30, 40, in color) for all segments separately and together; b) Sample CT values (mean as points, standard deviation as ranges) plotted against the sampling delay (days between symptom onset and sample collection).

To reveal the recent evolution of the three OROV genomic segments, we estimated three phylogenetic trees to determine their relationships with other isolates. Our analysis revealed that the novel OROV genome sequences sampled in 2024 clustered with sequences from the 2022-2024 epidemic into a monophyletic group, with strong bootstrap support (values of 1.0) across all three genomic segments (**Figure 3a-c**).

**Figure 3.**
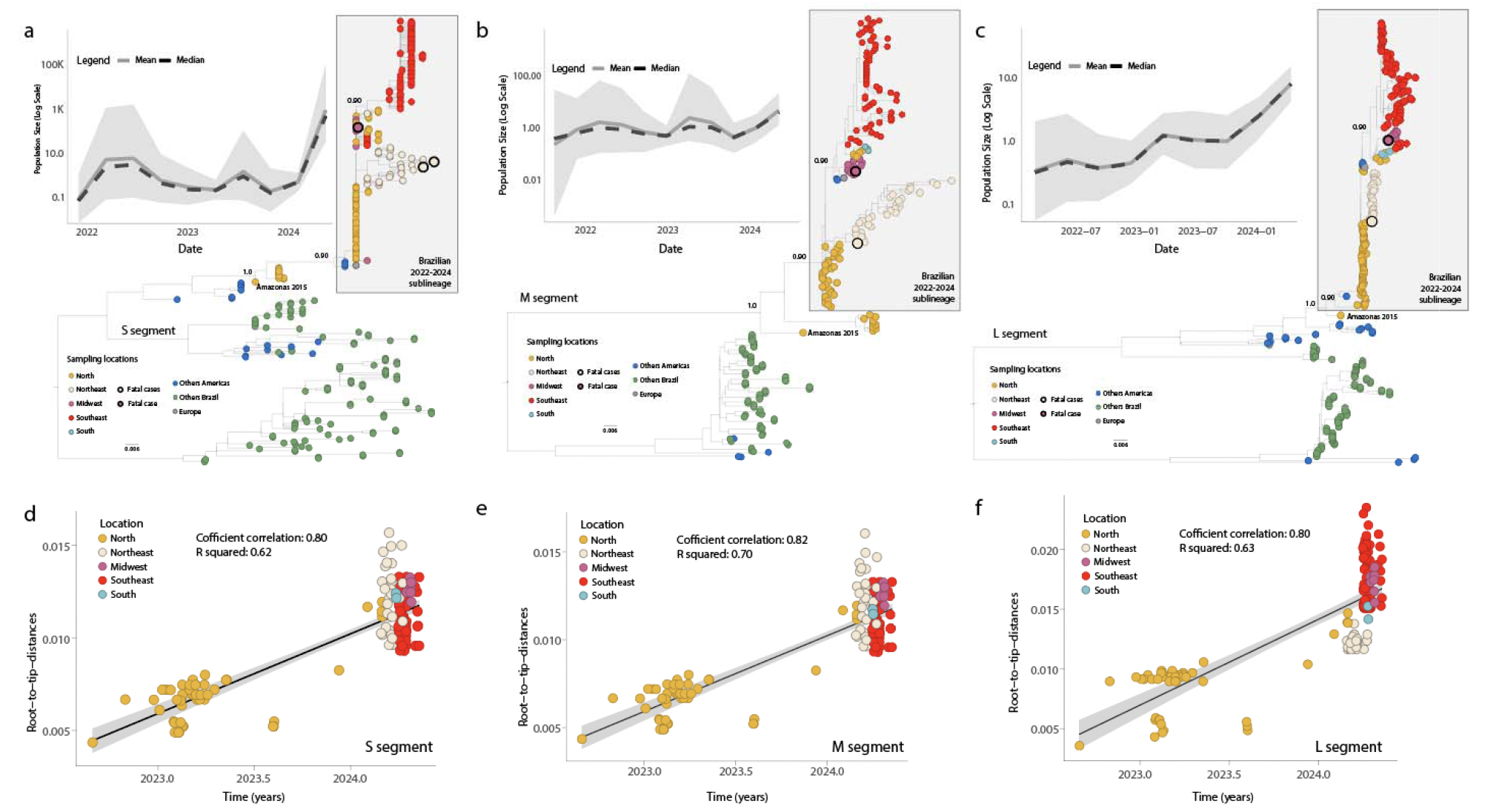
Molecular Evolution and Demographic History of OROV Segments S, M, and L. a-c) Maximum likelihood phylogenetic trees of the three OROV segments: S (n = 376), M (n = 231) (a), L (n = 303). Tips are color-coded according to the legend in the left corner; Inner plots indicate the effective population size (i.e., genetic diversity) of OROV infections (Log scale) over time estimated under the coalescent-based Bayesian Skygrid (BSKG) model (posterior median = solid lines, 95% HPD = pale areas) for each segment; d-f) Regression of sequence sampling dates against root-to-tip genetic distances in a maximum likelihood phylogeny of the Brazilian 2022-2024 expansion clade (n⍰=⍰254).

This clade included a basal sequence from the locality of Tefé, Amazonas state, sampled in 2015, indicating a likely Amazonian origin, and has rapidly expanded to other regions, following a north-to-south movement across the country **(Figure 3a-c)**. In addition, this lineage has a reassortant evolutionary history, possessing L and S segments from viruses found in Peru, Colombia, and Ecuador and an M segment from viruses detected in the eastern Amazon region. Detailed analysis of the epidemic expansion, based on estimates of effective population size, revealed a sharp increase at the beginning of 2024 **(Figure 3a-c, inner plots)**, coinciding with the timing of the national surge in OROV cases (**Figure 1)** (8). In addition, the phylogenetic trees for the S, M, and L segments (**Figure 3a-c**) revealed two main lineages. The first lineage appeared to be primarily composed of sequences from the northern and northeastern regions, while the second lineage was associated with the southeastern and southern regions. This geographical separation might reflect regional diversification of the virus as it spread outside the Amazon basin.

The observed topological discordance among the phylogenetic trees of different genomic segments indicated the likelihood of multiple reassortment or recombinant events, supporting the notion that reassortment/recombination is a common and potentially significant evolutionary mechanism in bunyaviruses. A recent preprint manuscript provides more information on the potential reassortment events on genomes from the Amazon Basin (9), so this will not be covered in detail in our manuscript.

To reconstruct viral movements across the country, we utilized smaller data sets derived from each genomic segment individually, focusing exclusively on the Brazilian 2022-2024 sublineage **(Figure 3 a-c)**. As there was a strong correlation between sampling time and root-to-tip divergence in all three data sets (**Figure 3 g-i**), we were able to use molecular clock models to infer evolutionary parameters. Accordingly, we estimated that the mean time of origin for this Brazilian 2022-2024 sublineage to be in early November 2021, with a 95% highest posterior density (HPD) interval ranging from early-August 2021 to early January 2022. This suggests continuous transmission within the Amazon basin (**Figure 4a-c**). Likely introduced in early 2015, this sublineage remained undetected due to insufficient active surveillance at the national level, initially spreading from the northern part of the country (Amazon basin) and subsequently moving towards the northeastern (Bahia state), midwestern (Mato Grosso state), southeastern (Minas Gerais state), and southern (Santa Catarina state) regions (**Figure 4a-c**). We expanded the data set related to this Brazilian 2022-2024 sublineage to include international genome sequences recently isolated in Peru and Italy. Analyses of these data revealed that OROV likely initially moved north to south within Brazil and then crossed borders to reach Peru (**Figure 4d**). Case reports recently detected in Italy are associated with returning travelers from Cuba, although genomes from the latter locality are currently unavailable. It is possible that OROV moved into Cuba sometime in between the events depicted in **Figure 4d** before being exported into Europe.

**Figure 4.**
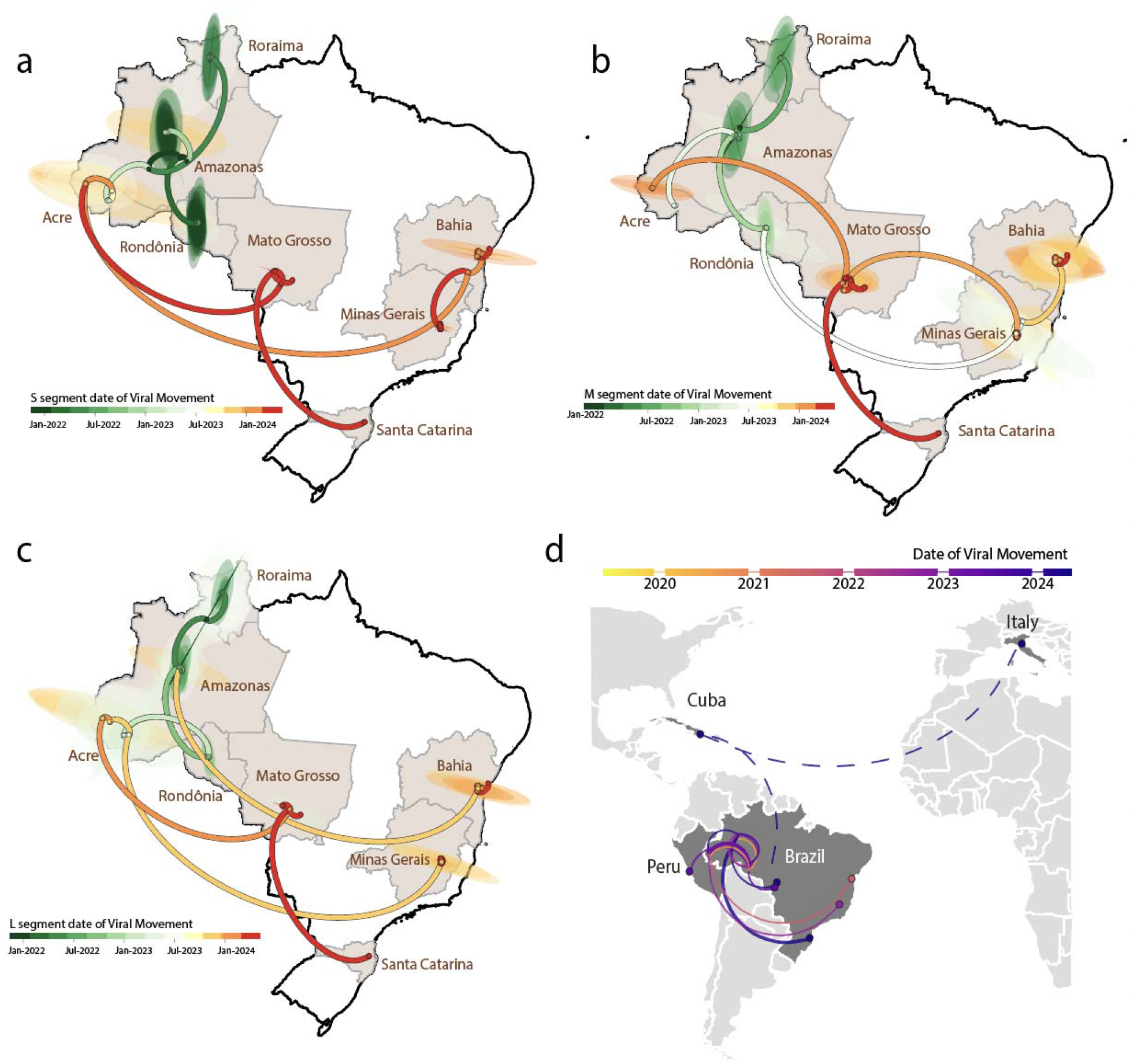
Inferred Viral Dissemination Patterns of OROV in Brazil and Globally. a-c) Phylogeographic reconstruction of the spread of OROV (segments S, M, and L) in Brazil. Circles represent nodes of the maximum clade credibility phylogeny and are colored according to their inferred time of occurrence. Shaded areas represent the 80% highest posterior density interval, depicting the uncertainty of the phylogeographic estimates for each node. Solid curved lines denote the links between nodes and the directionality of movement; d) Dissemination patterns of OROV within the Americas and Europe obtained from inferred ancestral-state reconstructions annotated and colored by region. The destination countries of viral exchange routes are shown with dots, with curves indicating the routes from the country of origin to the destination country in a counterclockwise direction. The dashed lines indicate the probable direction of dispersal concluded without evidence due to the paucity of genomes from Cuba.

## Discussion

The spread of the novel reassortant OROV lineage between 2022 and 2024 in the Brazilian Amazon region highlights the potential for this virus to emerge and the role of ecological and human factors in its dissemination (11, 12). Our study, along with recent findings (11, 12), underscores the importance of genomic surveillance and phylogenetic analysis in tracing the origin and movement of viral pathogens. The phylogenetic analyses from our study revealed that the OROV sequences sampled between 2022 and 2024 fell into a highly supported monophyletic group, with origins tracing back to a basal sequence from Tefé, Amazonas, Brazil, sampled in 2015. This indicates a likely Amazonian origin for this clade. These findings are consistent with the emergence of a novel reassortant lineage containing the M segment from viruses detected in the eastern Amazon region and the L and S segments from viruses found in Peru, Colombia, and Ecuador (10, 11). This reassortant lineage appears to have emerged in the central region of the Amazonas state between 2010 and 2014, demonstrating a long-range silent dispersion during the latter half of the 2010s.Our study identified approximately 21 reassortment events among the different genome segments, supporting the role of reassortment in the evolutionary history of OROV (11). It is possible that this contributed to the virus’s ability to adapt to new ecological niches and hosts, fueling its spread across different regions of Brazil. The sharp increase in OROV cases in different Brazilian regions along with the concurrent rise in real-time RT-PCR testing, reflects heightened surveillance and detection efforts underscoring the widespread concern and the proactive measures taken to monitor this emerging pathogen.

Our genome-based surveillance approach revealed a clear north-to-south movement of the virus within the country. The peak of OROV transmissions coincided with the rainy season in the Amazon basin, suggesting that environmental factors may play a major role in the virus’s transmission dynamics. Deforestation and climate change have contributed to the spread of OROV beyond the Amazon basin (32, 33). The alteration of ecosystems through deforestation disrupts natural habitats and promotes the migration of vectors, such as midges, into new areas (33). Climate change exacerbates this by altering weather patterns and creating favorable conditions for vector proliferation (34). These ecological changes facilitate the movement of OROV from its traditional Amazonian confines to other regions of Brazil and beyond. Recent studies have shown that increased deforestation rates and rising temperatures in the Amazon basin correlate with the expanded range of vector species capable of transmitting OROV (11, 35-40). This north-to-south dissemination pattern highlights the importance of addressing environmental and climate-related factors in controlling the spread of arboviruses. The prolonged cryptic circulation of the virus emphasizes the critical need for robust, active screening programs to monitor and control the spread of such pathogens effectively. The detection of three fatal cases associated with Oropouche virus (OROV) infection, even in patients without reported comorbidities, suggests that the clinical impact of OROV might be more significant than previously understood. This necessitates further investigations into adverse pregnancy outcomes and the potential for vertical transmission of OROV. On August 3rd, the Brazilian Ministry of Health (BrMoH) confirmed the first fetal death due to OROV with mother-to-child transmission in the state Pernambuco, located in the northeastern part of the country (41). This highlights the need for further research into the factors contributing to severe disease outcomes and the development of effective treatment strategies. In addition, the identification of long-range OROV migrations facilitated by human activities, as well as viral expansion beyond Brazilian borders to Peru, Cuba, and subsequently Europe, underscores the global implications of this emerging pathogen. The role of climate change in the expansion of vectors into new regions, coupled with increased human mobility, necessitates a coordinated international response to monitor and control the spread of OROV and other arboviruses.

Our study, in conjunction with recent research, emphasizes the critical need for continuous and widespread genomic surveillance. By understanding the evolutionary and epidemiological patterns of OROV, we can better anticipate and mitigate future outbreaks, ensuring more effective public health responses both within Brazil and globally. This approach will be instrumental in managing the ongoing spread of OROV and preparing for potential future threats posed by similar viral pathogens.

## Data Availability

NA

## Author Contributions

Conceptualization: V.F., J.L., T.d.O., L.C.J.A., and M.G.; Methodology: F.C.M.I., F.M.P., E.C.O., J.T.N.R., M.H.M., V.F., T.E.R.A., N.R.G., L.M.R.T., M.K.A.G., V.B.M., A.A.R., A.R., A.G.F., A.L.S.M., B.M.N.F., C.F.C.A., D.S.P., F.G.M.L., F.V.M.S., G.C.P., H.T., J.D.,P.,C.A., K.M.F.M., K.R.V., L.C.S., L.C.M.S., L.C.V.F., L.O.M., M.A.C., M.S.O., M.P.D.A., M.C., M.T.L., M.A.P., M.L.C.R., P.E.S.S., P.R., R.S.A., R.S., R.T.G., R.V.C., S.G., S.C.F.S., S.A.A.B., S.H.S.P.P., S.I.R.A., S.F.S., W.V.V., D.P.M., E.C.H., T.d.O., J.L., L.C.J.A., and M.G.; Investigation: F.C.M.I., F.M.P., E.C.O., J.T.N.R., M.H.M., V.F., T.E.R.A., N.R.G., L.M.R.T., M.K.A.G., V.B.M., A.A.R., A.R., A.G.F., A.L.S.M., B.M.N.F., C.F.C.A., D.S.P., F.G.M.L., F.V.M.S., G.C.P., H.T., J.D.,P.,C.A., K.M.F.M., K.R.V., L.C.S., L.C.M.S., L.C.V.F., L.O.M., M.A.C., M.S.O., M.P.D.A., M.C., M.T.L., M.A.P., M.L.C.R., P.E.S.S., P.R., R.S.A., R.S., R.T.G., R.V.C., S.G., S.C.F.S., S.A.A.B., S.H.S.P.P., S.I.R.A., S.F.S., W.V.V., D.P.M., E.C.H., T.d.O., J.L., L.C.J.A., and M.G.; Data curation: V.F., J.L., and M.G.; Original draft preparation: J.L., M.G.; Review and editing: F.C.M.I., F.M.P., E.C.O., J.T.N.R., M.H.M., V.F., T.E.R.A., N.R.G., L.M.R.T., M.K.A.G., V.B.M., A.A.R., A.R., A.G.F., A.L.S.M., B.M.N.F., C.F.C.A., D.S.P., F.G.M.L., F.V.M.S., G.C.P., H.T., J.D.,P.,C.A., K.M.F.M., K.R.V., L.C.S., L.C.M.S., L.C.V.F., L.O.M., M.A.C., M.S.O., M.P.D.A., M.C., M.T.L., M.A.P., M.L.C.R., P.E.S.S., P.R., R.S.A., R.S., R.T.G., R.V.C., S.G., S.C.F.S., S.A.A.B., S.H.S.P.P., S.I.R.A., S.F.S., W.V.V., D.P.M., E.C.H., T.d.O., J.L., L.C.J.A., and M.G.; Visualization: V.F., J.L., and M.G. All authors have read and agreed to the published version of the manuscript.

## Acknowledgments

This study was supported by the National Institutes of Health USA grant U01 AI151698 for the United World Arbovirus Research Network (UWARN), the CRP-ICGEB RESEARCH GRANT 2020 Project CRP/BRA20-03, Contract CRP/20/03, and the Rede Unificada de Análises Integradas de Arbovírus de Minas Gerais (REDE UAI-ARBO-MG), financed by Fundação de Amparo à Pesquisa do Estado de Minas Gerais (FAPEMIG), grant number RED-00234-23. M. Giovanetti’s funding is provided by PON “Ricerca e Innovazione’’ 2014-2020. T.E.R.A. is supported by Conselho Nacional de Desenvolvimento Científico e Tecnológico (CNPq) under the process number 153597/2024-0. F.C.M.I. is supported by FAPEMIG under process number BIP-00123-23. The authors would also like to acknowledge the Global Consortium to Identify and Control Epidemics – CLIMADE (https://climade.health/).

## Supplementary figures

**Figure S1.**
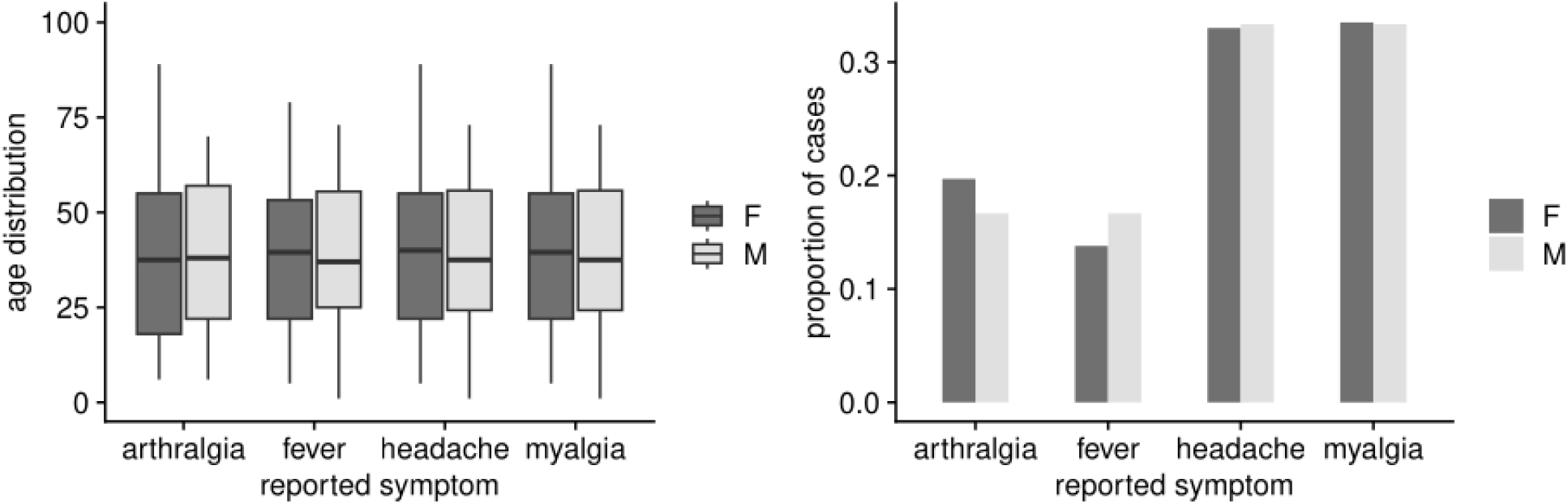
Symptoms versus age and gender. The (left) age distribution of cases reported with different symptoms and (b) proportion of cases reporting different symptoms, both disaggregated by gender. Data includes the metadata for the sequenced samples.

**Figure S2.**
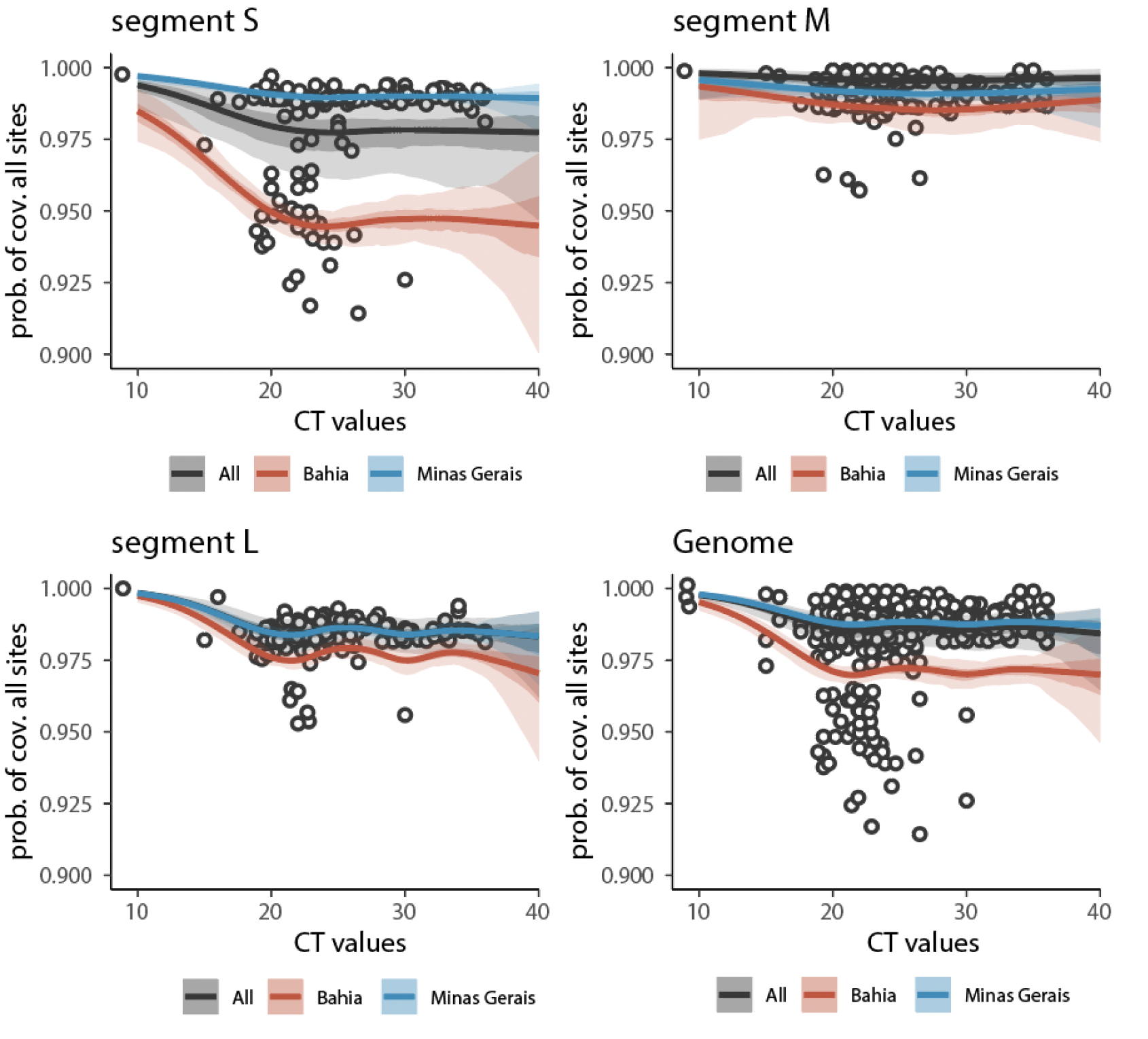
Sequence Coverage Probability Across Genome Segments in Different Brazilian States. The figure displays the probability of coverage for all sites across four different genome segments (S, M, L, and the entire genome) in relation to the CT value. The probability was estimated using a Generalized Additive Model (GAM; as done previously e.g. for CHIKV sequencing data [Giovanetti et al Emerg Infect Dis. 2023;29(9):1859-1863]). The GAM was defined with a binomial family, with coverage modeled as the proportion of sites recovered from sequencing, using sample state (Brazilian state) as random effect. Solutions presented (in color) include the specific ones for the two states with most sequencing (Bahia, red; Minas Gerais, blue) and the general model output (gray).

